# The impact of traffic isolation in Wuhan on the spread of 2019-nCov

**DOI:** 10.1101/2020.02.04.20020438

**Authors:** Gehui Jin, Jiayu Yu, Liyuan Han, Shiwei Duan

**Affiliations:** Ningbo University

## Abstract

The 2019-nCoV outbreak occurred near the Chinese Spring Festival transport period in Wuhan. As an important transportation center, the migration of Wuhan accelerated the spread of 2019-nCoV across mainland China. Based on the cumulative Baidu migration index (CBMI), we first analyzed the proportion of Wuhan’s migrant population to other cities. Our results confirm that there is a significant correlation between the export population of Wuhan and reported cases in various regions. We subsequently found that the mortality rate in Hubei Province was much higher than that in other regions of mainland China, while the investigation of potential cases in Wuhan was far behind other provinces in Mainland China, which indicates the effectiveness of early isolation.

---

2019-nCoV outbreak occurred near the Chinese Spring Festival transport period (January 10 to February 8, 2020) in Wuhan. As an important transportation center, the migration of Wuhan accelerated the wide spread of 2019-nCoV across mainland China. From 10:00 on January 23, 2020, the transportation in Wuhan was suspended^1^ (Figure 1a). Although the closure of the city has been implemented, more than 5 million people have left Wuhan in the recent days^2^.

Based on cumulative Baidu migration index (CBMI)^3^ from January 16 to January 25, 2020, we found that 74.1% of Wuhan’s migrant population went to other cities in Hubei, 5.6% Wuhan’s migrant population has entered Henan, and other provinces account for a small proportion (Figure 1b). In Hubei Province, most migrant population from Wuhan entered Xiaogan (20%), Huanggang (19%), and Jingzhou (10%) (Figure 1c).

**Figure 1:**
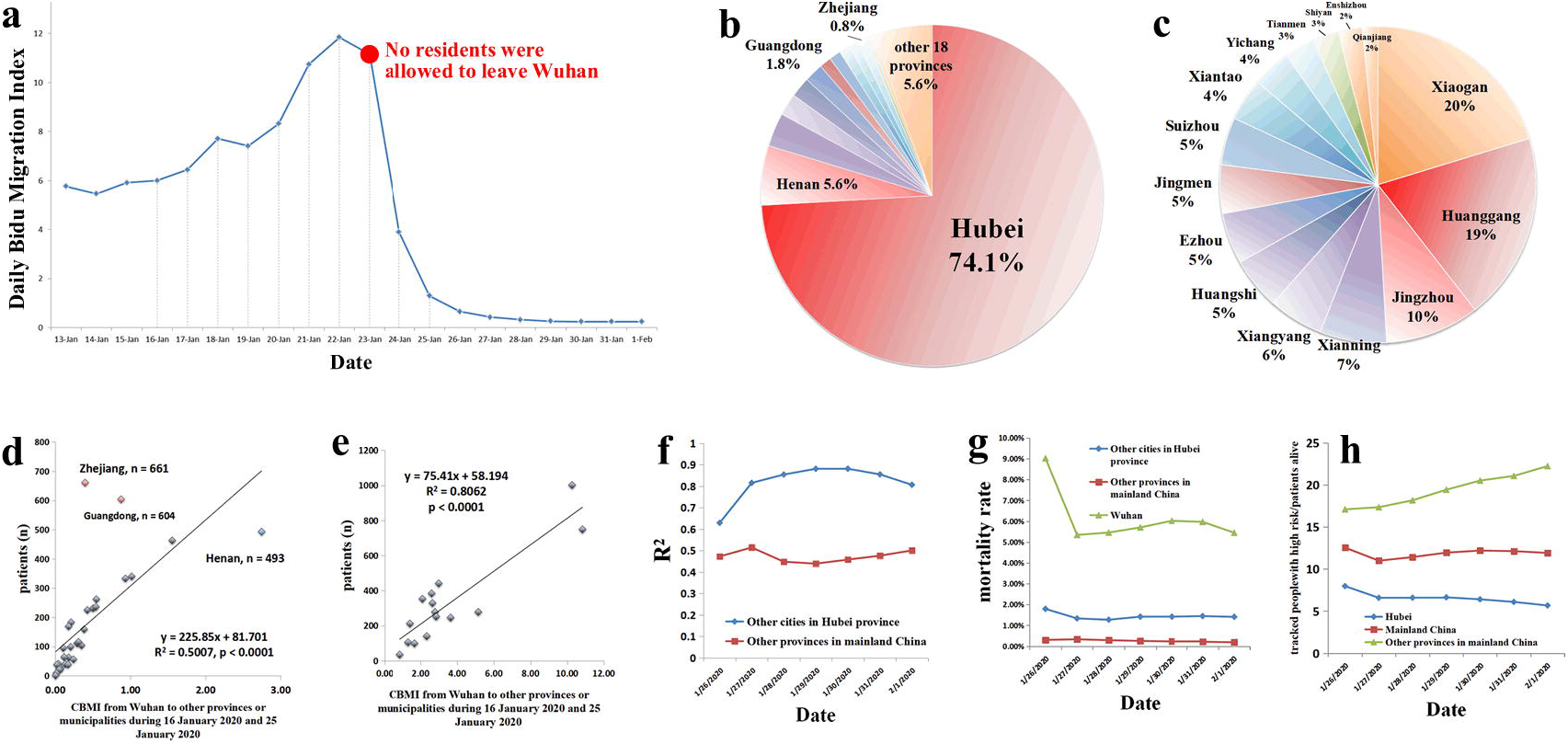
The impact of traffic isolation in Wuhan on the spread of 2019-nCov CBMI: cumulative Baidu outbound migration index. Figure 1a: The daily Baidu outbound migration index of Wuhan from January 13 to February, 2020. Figure 1b: The distribution of cumulative Baidu outbound migration index (CBMI) of Wuhan from January 16 to January 25, 2020 in mainland China. Figure 1c: The distribution of Wuhan CBMI (from January 16 to January 25, 2020) in Hubei province. Figure 1d: The correlation of reported patients (February 1, 2020) in other non-Hubei provinces of mainland China with Wuhan CBMI (from January 16 to January 25, 2020). Figure 1e: The correlation of reported patients (February 1, 2020) in other non-Wuhan cities of Hubei province with Wuhan CBMI (from January 16 to January 25, 2020). Figure 1f: The R2 values in the correlation tests between reported patients (from January 26 to February 1, 2020) and Wuhan CBMI (from January 16 to January 25, 2020). Figure 1g: The mortality rates in Wuhan, other cities in Hubei province, and other provinces in mainland China. Figure 1h: The ratios of the tracked high-risk people to patients alive (from January 26 to February 1, 2020) in Hubei province, other provinces, and mainland China.

We used 2019-nCoV-related data published by the National and Provincial Health and Health Committees from January 20 to February 1, 2020 and analyzed the correlation between the CBMI of Wuhan and the 2019-nCoV cases by linear regression test.

We analyzed the correlation between the number of 2019-nCoV related pneumonia cases and the CBMI of Wuhan (Figures 1d-1e), and found that the number of reported cases in the other non-Hubei regions of mainland China was highly correlated with the CBMI of Wuhan (Figure 1d, R^2^ = 0.50 p <0.0001). In addition, the number of reported cases in other non-Wuhan cities in Hubei Province (February 1 of 2020) was also highly correlated with the CBMI of Wuhan during January 16 and 25 of 2020 (Figure 1e, R^2^ = 0.81, p <0.0001). We further found that the correlation between Wuhan CBMI and reported cases was very stable in other non-Hubei regions of mainland China (Figure 1f, R^2^ = 0.43-0.51) and other non-Wuhan cities in Hubei Province (Figure 1f, R^2^ = 0.62-0.88). Our results confirm that there is a significant correlation between the export population of Wuhan and reported cases in various regions.

In Figure 1d, we also observed that the reported cases are relatively small, although Wuhan CBMI in Henan Province is large (5.6%, Figure 1b). We speculate that Henan Province’s earlier intervention measures (such as traffic control and designated hospitals) effectively reduced the proliferation of 2019-nCov^4^.

In addition, the total number of reported cases in Zhejiang Province was higher than expected (Figure 1d). In Zhejiang Province, Wenzhou was the city most frequently traded with Wuhan, had the highest number of reported cases. From January 19th to January 23rd, 2020, Wenzhou’s Wuhan CBMI (0.11) ranked first in Zhejiang Province. We speculated that some patients returned to Wenzhou before Wuhan closed the city, resulting in the widespread propagation of 2019-nCoV in Wenzhou. Similarly, Guangdong Province and Wuhan had close economic ties and personnel exchanges, leading to a higher risk of infection with 2019-nCoV.

Subsequently, we further analyzed the mortality rate of 2019-nCoV pneumonia in different regions (Figure 1g). We found that the mortality rate in Hubei Province (3.24%; Wuhan: 5.45%) was much higher than that in other regions of mainland China (0.19%). We have also found that the investigation of potential cases in Wuhan was far behind other provinces in Mainland China (Figure 1h). We speculated that this might be related to a large number of patients, limited medical resources, and the lack of manpower to track suspected cases.

From a public health perspective, effective isolation and reduced exposure to patients who have been diagnosed are the most important. Our results confirm that the mandatory traffic segregation in Wuhan has greatly reduced the nationwide spread of 2019-nCov. At the same time, our results also show that the export of the population in Wuhan before the closure of the city will lead to the spread of severe epidemics in other cities in Hubei.

## Data Availability

All the data are from the public resources.

## Acknowledgement

This work is supported by the K.C. Wong Magna Fund in Ningbo University.

